# Development and validation of multivariable prediction models for adverse COVID-19 outcomes in IBD patients

**DOI:** 10.1101/2021.01.15.21249889

**Authors:** John Sperger, Kushal S. Shah, Minxin Lu, Xian Zhang, Ryan C. Ungaro, Erica J. Brenner, Manasi Agrawal, Jean-Frederic Colombel, Michael D. Kappelman, Michael R. Kosorok

**Affiliations:** Department of Biostatistics, University of North Carolina, Chapel Hill, NC; Division of Pediatric Gastroenterology, Department of Pediatrics, University of North Carolina, Chapel Hill, NC; The Henry D. Janowitz Division of Gastroenterology, Icahn School of Medicine at Mount Sinai, NY

## Abstract

**Importance:** Risk calculators can facilitate shared medical decision-making^1^. Demographics, comorbidities, medication use, geographic region, and other factors may increase the risk for COVID-19-related complications among patients with IBD^2,3^.

**Objectives:** Develop an individualized prognostic risk prediction tool for predicting the probability of adverse COVID-19 outcomes in patients with IBD.

**Design, Setting, and Participants:** This study developed and validated prognostic penalized logistic regression models^4^ using reports to Surveillance Epidemiology of Coronavirus Under Research Exclusion for Inflammatory Bowel Disease (SECURE-IBD) from March–October 2020. Model development was done using a training data set (85% of cases reported March 13 – September 15, 2020), and model validation was conducted using a test data set (the remaining 15% of cases plus all cases reported September 16–October 20, 2020).

**Main Outcomes and Measures:** COVID-19 related:

1. Hospitalization+: composite outcome of hospitalization, ICU admission, mechanical ventilation, or death
2. ICU+: composite outcome of ICU admission, mechanical ventilation, or death
3. Death

We assessed the resulting models’ discrimination using the area under the curve (AUC) of the receiver-operator characteristic (ROC) curves and reported the corresponding 95% confidence intervals (CIs).

**Results:** We included 2709 cases from 59 countries (mean age 41.2 years [s.d. 18], 50.2% male). A total of 633 (24%) were hospitalized, 137 (5%) were admitted to the ICU or intubated, and 69 (3%) died. 2009 patients comprised the training set and 700 the test set.

The models demonstrated excellent discrimination, with a test set AUC (95% CI) of 0.79 (0.75, 0.83) for Hospitalization+, 0.88 (0.82, 0.95) for ICU+, and 0.94 (0.89, 0.99) for Death. Age, comorbidities, corticosteroid use, and male gender were associated with higher risk of death, while use of biologic therapies was associated with a lower risk.

**Conclusions and Relevance:** Prognostic models can effectively predict who is at higher risk for COVID-19-related adverse outcomes in a population of IBD patients. A free online risk calculator (https://covidibd.org/covid-19-risk-calculator/) is available for healthcare providers to facilitate discussion of risks due to COVID-19 with IBD patients. The tool numerically and visually summarizes the patient’s probabilities of adverse outcomes and associated CIs. Helping physicians identify their highest-risk patients will be important in the coming months as cases rise in the US and worldwide. This tool can also serve as a model for risk stratification in other chronic diseases.

**Key Points:** *Question:* How well can a multivariable risk model predict the risk of hospitalization, intensive care unit (ICU) stay, or death due to COVID-19 in patients with inflammatory bowel disease (IBD)?

*Findings:* Multivariable prediction models developed using data from an international voluntary registry of IBD patients and available for use online (https://covidibd.org/) have very good discrimination for predicting hospitalization (Test set AUC 0.79) and excellent discrimination for ICU admission (Test set AUC 0.88) and death (Test set AUC 0.94). The models were developed with a training sample of 2009 cases and validated in an independent test sample of 700 cases comprised of a random sub-sample of cases and all cases entered in the registry during a one-month period after model development.

*Meaning:* This risk prediction model may serve as an effective tool for healthcare providers to facilitate conversations about COVID-19-related risks with IBD patients.

## Introduction

Since the onset of the novel coronavirus disease 2019 (COVID-19) pandemic, almost 50 million cases have been reported globally. Many countries, including the United States (US), are reporting record numbers of new cases as of November 2020^5^. While the majority of cases are mild, patients with at least one comorbidity are at higher risk of adverse outcomes, including hospitalization, respiratory failure, or death^6,7^. Risk calculators can facilitate shared decision making between patients and healthcare providers^1^, and such tools have been created to predict death due to COVID-19 in US patients 65 years and older^8^, to determine hospitalization risk^9^, and to guide early vaccine allocation. Patients with inflammatory bowel disease (IBD) are prescribed immunosuppressive medications such as corticosteroids, immunomodulators, biologic therapies, and Janus-kinase inhibitors, which are linked with a higher risk of viral infection^10,11^. To help healthcare providers and patients navigate these myriad potential risk factors, we developed and validated penalized multivariable logistic regression models for predicting the probability of hospitalization, intensive care unit (ICU) admission, and death due to COVID-19 in patients with IBD. We utilized an international registry of 2,709 IBD patients with COVID-19 from 59 countries. We also developed a free, publicly available personalized risk calculator utilizing the final models that is available online (https://covidibd.org/covid-19-risk-calculator/). Reporting follows Transparent reporting of a multivariable prediction model for individual prognosis or diagnosis (TRIPOD)^12^ guidelines.

## Methods

### Source of Data

The Surveillance Epidemiology of Coronavirus Under Research Exclusion for Inflammatory Bowel Disease (SECURE-IBD) database (www.covidibd.org) is an international registry to study outcomes of COVID-19 in pediatric and adult IBD patients^13^. SECURE-IBD is a voluntary registry with ongoing data collection where healthcare providers can report cases of COVID-19 in IBD patients, confirmed by polymerase chain reaction (PCR) or antibody testing. Healthcare providers are instructed to report cases of severe outcomes after a minimum of seven days from the onset of symptoms and after a sufficient time has passed to observe the disease course through the resolution of acute illness or death. In the event that a patient’s status changed after submission, reporters are instructed to re-report and contact the research team. Reporters were not explicitly informed of what data could be used as predictors or outcomes, but being a voluntary registry, reporters were not blinded. A fuller account of the data collection is given in Brenner et al. (2020)^14^.

### Participants

We included all patients reported to the registry from March 13, 2020, the data collection start date, through October 20, 2020. For model development, a training sample consisting of 85% of the entire surveillance data set available as of September 15, 2020 was used. The random split was done using stratified random sampling based on an ordinal version of the outcome. The test data set consisted of the remaining 15% of the data available on September 15, plus all of the additional cases reported to the registry between September 16, 2020 and October 20, 2020.

We reported means and standard deviations for continuous variables, counts for categorical variables, and proportions for binary variables. We reported the missing data for all variables. We did not include P values in our descriptive tables following the Strengthening the Reporting of Observational Studies in Epidemiology (STROBE) guidelines^15^.

### Outcomes

We examined three primary outcomes: 1) hospitalization or death (Hospitalization+), 2) ICU admission, mechanical ventilation or death (ICU+), and 3) death due to COVID-19 related causes (Death). Patients may experience multiple outcomes. All outcomes were reported by the patient’s healthcare provider at the time of the case report.

### Predictors

As our aim was to create models and a risk stratification tool intended to allow physicians to inform patients of their risk before presenting with COVID-19, we restricted our attention to predictors that would be available during a routine consultation. COVID-19 presenting symptoms and information about the COVID-19 treatment received were therefore not included in this analysis. All predictors were reported by the patients’ healthcare provider.

A full description of the predictors is available in **Supplement document eTable 1**. Demographic predictors included age, country of residence, state of residence (for US cases), gender, race, and ethnicity. Racial indicators included White, Black, and Asian. American Indian and Pacific Islander indicators were excluded due to low prevalence. Multi-racial patients belong to multiple categories. As only one patient had reported a gender other than male or female, only two genders were considered in the analysis. Due to the nature of reporting, ethnicity, gender, and race should be interpreted as provider-perceived race and gender. Assessing race and ethnicity is important for identifying potential health inequities in COVID-19 related outcomes. For cases from US states with very low prevalence in the registry, a more general geographic predictor (census region or census division) was used in place of the state itself. Clinical predictors included height, weight, body mass index (BMI, study derived), IBD diagnosis [Crohn’s disease (CD), ulcerative colitis (UC), or IBD unspecified (IBDU)], and IBD disease activity as defined by physician global assessment. We also included IBD medication indicators for overall drug category (e.g., biologic therapy) and subcategory [e.g., anti-tumor necrosis factor (TNF)] at the time of COVID-19 diagnosis. Additionally, dosage information was included for prednisone, 6-mercaptopurine, and azathioprine.

**Table 1.** Main Characteristics of COVID-19 IBD Patients in the Study^a^.

| Characteristics | Training Data<br>(n=2009) | Test Data<br>(n=700) | Overall<br>(n=2709) |
| --- | --- | --- | --- |
| Age, mean (SD), years | 42.2 (18.2) | 38.7 (17.4) | 41.2 (18.0) |
| Gender, No. (%) |  |  |  |
| Female | 982 (48.9%) | 344 (49.1%) | 1326 (48.9%) |
| Male | 998 (49.7%) | 341 (48.7%) | 1339 (49.4%) |
| Other | 1 (0.0%) | 0 (0.0%) | 1 (0.0%) |
| Asian<sup>b</sup>, No. (%) | 112 (5.6%) | 38 (5.4%) | 150 (5.5%) |
| Black<sup>b</sup>, No. (%) | 138 (6.9%) | 39 (5.6%) | 177 (6.5%) |
| White<sup>b</sup>, No. (%) | 1603 (79.8%) | 547 (78.1%) | 2150 (79.4%) |
| Hispanic/Latino, No. (%) | 350 (17.4%) | 115 (16.4%) | 465 (17.2%) |
| Missing | 375 (18.7%) | 120 (17.1%) | 495 (18.3%) |
| BMI, mean (SD) | 26.0 (6.50) | 25.2 (6.26) | 25.8 (6.44) |
| Missing | 398 (19.8%) | 106 (15.1%) | 504 (18.6%) |
| Current smoker, No. (%) | 61 (3.0%) | 25 (3.6%) | 86 (3.2%) |
| Disease type, No. (%) |  |  |  |
| Crohn's disease | 1115 (55.5%) | 401 (57.3%) | 1516 (56.0%) |
| Ulcerative colitis | 854 (42.5%) | 278 (39.7%) | 1132 (41.8%) |
| Cardiovascular disease, No. (%) | 133 (6.6%) | 43 (6.1%) | 176 (6.5%) |
| Diabetes, No. (%) | 117 (5.8%) | 30 (4.3%) | 147 (5.4%) |
| Hypertension, No. (%) | 243 (12.1%) | 67 (9.6%) | 310 (11.4%) |
| Count of comorbidities, mean (SD) | 0.544 (0.938) | 0.479 (0.879) | 0.527 (0.923) |
| Biologic therapy, No. (%) | 1203 (59.9%) | 437 (62.4%) | 1640 (60.5%) |
| Tumor Necrosis Factor inhibitor, No. (%) | 796 (39.6%) | 313 (44.7%) | 1109 (40.9%) |
| Anti-integrin, No. (%) | 213 (10.6%) | 72 (10.3%) | 285 (10.5%) |
| IL 12/23 inhibitor, No. (%) | 187 (9.3%) | 47 (6.7%) | 234 (8.6%) |
| 5-Aminosalicylates, No. (%) | 636 (31.7%) | 200 (28.6%) | 836 (30.9%) |
| Sulfasalazine, No. (%) | 64 (3.2%) | 24 (3.4%) | 88 (3.2%) |
| Mesalamine, No. (%) | 561 (27.9%) | 175 (25.0%) | 736 (27.2%) |
| Immunomodulators, No. (%) | 459 (22.8%) | 140 (20.0%) | 599 (22.1%) |
| Methotrexate, No. (%) | 81 (4.0%) | 24 (3.4%) | 105 (3.9%) |
| Azathioprine or 6-Mercaptopurine, No. (%) | 367 (18.3%) | 110 (15.7%) | 477 (17.6%) |
| Corticosteroids, No. (%) | 209 (10.4%) | 65 (9.3%) | 274 (10.1%) |
| Budesonide, No. (%) | 59 (2.9%) | 17 (2.4%) | 76 (2.8%) |
| Oral or parenteral steroids, No. (%) | 154 (7.7%) | 49 (7.0%) | 203 (7.5%) |
| Janus kinase inhibitors (tofacitinib), No. (%) | 30 (1.5%) | 9 (1.3%) | 39 (1.4%) |
| Hospitalization+, No. (%) | 499 (24.8%) | 138 (19.7%) | 637 (23.5%) |
| Missing | 37 (1.8%) | 17 (2.4%) | 54 (2.0%) |
| ICU+, No. (%) | 133 (6.6%) | 36 (5.1%) | 169 (6.2%) |
| Missing | 49 (2.4%) | 22 (3.1%) | 71 (2.6%) |
| Death, No. (%) | 57 (2.8%) | 12 (1.7%) | 69 (2.5%) |
| Missing | 33 (1.6%) | 14 (2.0%) | 47 (1.7%) |

<sup>a</sup> Missingness is only reported for predictors with >5% missing values, and for all outcomes. Only comorbidities with an overall prevalence above 5% are shown in this table. For a complete list of characteristics and number and percentage of missing values, see supplement eTable 2.
<sup>b</sup> Individuals can be assigned to more than one physician-reported racial group (White, Black, Asian).

For categorical (including binary) predictors without a meaningful reference level, all levels were included in the model. Quadratic terms were considered for all continuous covariates. Interactions were considered based on a combination of subject matter expert advice and a minimum threshold of thirty observations for every cell for interactions involving two binary predictors.

### Missing Data

Multiple imputation^16^ of the covariates and outcomes was performed using multivariate imputation by chained equations (MICE)^17^ to address missing data. A total of thirty imputed data sets were created. Imputation was performed separately on the training and test data to prevent inducing dependence between the training and the test data through the imputation models. For transformed variables that are derived from other covariates (e.g., BMI from height and weight), we imputed the missing root variables and then created the transformed variable to ensure that the relationship between the transformed variable and its inputs was preserved.

Table 1 includes the level of missing data in each of the covariates included in the analysis. Medication variables, clinical descriptions of disease and severity, location, age, and gender all had very low levels of missingness, ranging from zero to under five percent. There were three covariates with a moderate amount of missing data — height, weight, and ethnicity were missing in approximately 20% of patients.

### Statistical Analysis

The 10-fold cross-validation deviance averaged across each of the imputed data sets was used to decide between LASSO, Ridge, or Elastic net penalties and to choose the value of the regularization parameter^4,18^. Separate logistic regression models were fit for each of the outcomes. Smoothing splines for continuous covariates^19^, a multinomial model, Group LASSO^20^, and Sparse Group LASSO^21^ were all investigated as potential methods, but the performance improvement in terms of the cross-validation deviance was not sufficient to justify the additional complexity and computational time. The nonparametric resampling bootstrap was used to generate 1,000 samples, and for each bootstrap sample, the same sampled participants were used across the 30 imputed data sets^22^. A total of 30,000 (30 × 1000) fitted models were created. Predicted probabilities were created by averaging the predictions from all models. We used the sample mean of the bootstrap distribution of the predicted probabilities for the final predicted probability and the percentiles of the bootstrap distribution to find the 90% confidence interval for the risk estimate. Risk groups were not created.

To assess the performance of the resulting predictions, we created Receiver Operator Characteristic (ROC) curves and calculated the corresponding area under the curve (AUC) using the held-out test data set for the imputed data sets^22^. We provide two graphical summaries of the resulting models: 1) a summary of the sign distribution showing, across bootstrap replications and imputed data sets, the proportion of estimated associations which were negative (better outcomes), zero, or positive, and 2) box plots of the estimated effect on the log-odds scale. We opted to show box plots instead of confidence intervals to highlight the exploratory nature of the results for individual predictors. Lasso estimates are biased, and the lack of a priori hypotheses makes statistical significance testing inappropriate. We first defined a set of contrasts in order to make meaningful comparisons while accounting for the second-order terms in the model rather than report results for every parameter. The contrast matrices are available on a public repository https://github.com/KosorokLab/CovidIBDRiskCalc in a CSV format.

Predictions are averaged over bootstrap replications and imputed data sets, and so there is no single set of model coefficients to report. Because the logistic link function is non-linear, the predicted probability from averaging over predictions from each fitted model does not equal the predicted probability from averaging over coefficients across the models. Additionally, averaging the model coefficients would result in none of the coefficients being equal to zero unless that coefficient is equal to zero in every fitted model. Instead of reporting a misleading model summary, we opted to make all the model coefficients available online (https://github.com/KosorokLab/CovidIBDRiskCalc).

### Software

The analysis was conducted using R^23^ version 4.0.2 and the tidyverse^24^, glmnet^25^, glmnetUtils^26^, mice^17^, magrittr^27^, future^28^, and pROC^29^ packages. The online calculator was created using shiny^30^. The most recent draw for the Carolina Pick 4 lottery (https://nclottery.com/Pick4) at the time of analysis was used for the random number generation seed. The code used to conduct the analysis is available on GitHub (https://github.com/KosorokLab/CovidIBDRiskCalc). This does not include the study data, but the estimated model coefficients are available.

## Results

### Participants

A total of 2,709 patients were reported to the registry, split into a training set of 2009 patients and a test data set of 700 patients. The test data set was comprised of 366 from the 15% split and 334 patients added to the registry after model fitting and before manuscript submission. Table 1 provides demographic, clinical, medication, and outcome descriptive summaries for the training set, test set, and the whole sample. A total of 633 (24%) patients were hospitalized, 137 (5%) were admitted to the ICU or intubated, and 69 (3%) patients died. The cohort has 1076 (40%) patients from the United States, with the rest coming from a variety of other countries summarized in **Table 1**.

### Model Performance

The models have excellent discrimination, with an AUC and associated 95% confidence interval estimated on the test data set averaged over the imputations of 0.79 (0.75, 0.83) for Hospitalization+, 0.88 (0.82, 0.95) for ICU+, and 0.94 (0.89, 0.99) for death.

### Predictors of hospitalization, intensive care, and death

Figures 1 & 2 show the estimated coefficient sign distribution and the effect on the log-odds scale for the ten contrasts most strongly associated with each outcome respectively. Consistent with other studies^7,31^ on risk factors for hospitalization and death, we find older age, male gender, and comorbidities to be associated with worse outcomes due to COVID-19. White race is associated with a lower risk of Hospitalization+ in 89.2% of our replications but is not consistently selected in the models for ICU+ (30%) or death (10%). These plots, not restricted to the top ten effects, are available in the supplement for all demographic, clinical, and medication predictors (eFigures 1 & 2), for countries (eFigures 3 & 4), and for US regions (eFigures 5 & 6).

**Figure 1:**
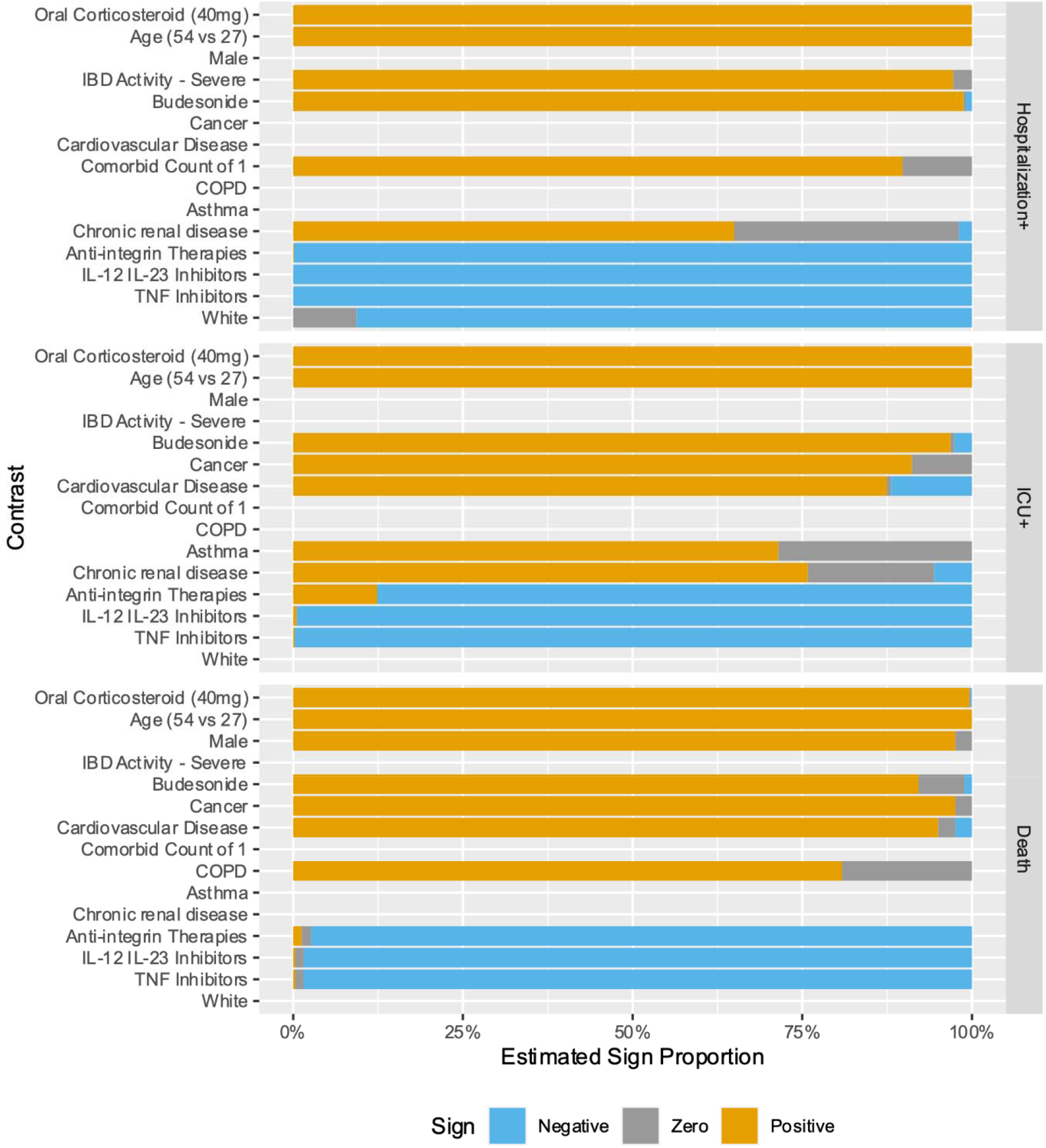
Estimated Contrast Sign Distribution

**Figure 2:**
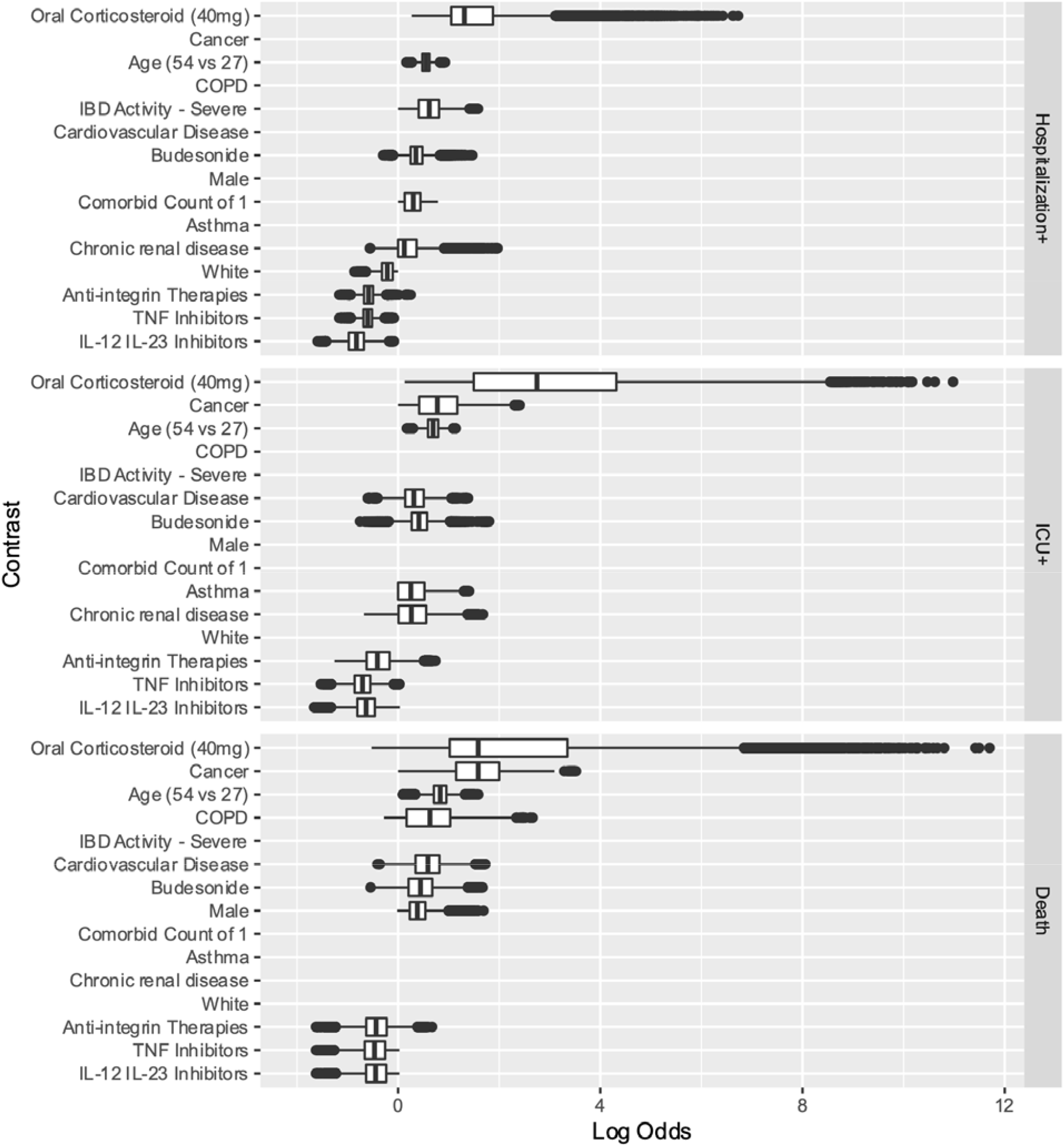
Estimated Contrast Effect Size Distribution

Corticosteroids are associated with a higher risk of Hospitalization+, ICU+, and death. Oral corticosteroid use is the most important predictor, in terms of the magnitude of the absolute value of the coefficient, for Hospitalization+, ICU+, and death (Figure 2). Biologic medicines are associated with a lower risk of Hospitalization+, ICU+, and death, with integrin antagonists having directionally smaller effects than tumor necrosis factor (TNF) antagonists or interleukin (IL) 12/23 inhibitors.

### Online Risk Tool

The online risk calculator where physicians can enter their patient’s information and receive predictions from our models is freely available at (http://shiny.bios.unc.edu/secure-ibd-risk-calc/). The SECURE-IBD COVID-19 Risk Calculator was designed for physicians to use during consultations with their patients and includes detailed clinical characteristics, including demographics, disease diagnosis information, comorbidities, and current medications. Daily dosage may optionally be entered for certain medications. The output of the risk calculator numerically and visually summarizes the patient’s probabilities of adverse outcomes and associated prediction intervals among the three nested outcomes discussed earlier. Figure 3 displays the results for two example patients and their associated probabilities (and 90% confidence intervals) of adverse outcomes if they were to contract COVID-19. The interactive application could provide a reliable basis for distinguishing between high- and low-risk patients to aid in personalizing clinical guidance on decisions about precautions, returning to normal activities, and vaccination.

**Figure 3A.**
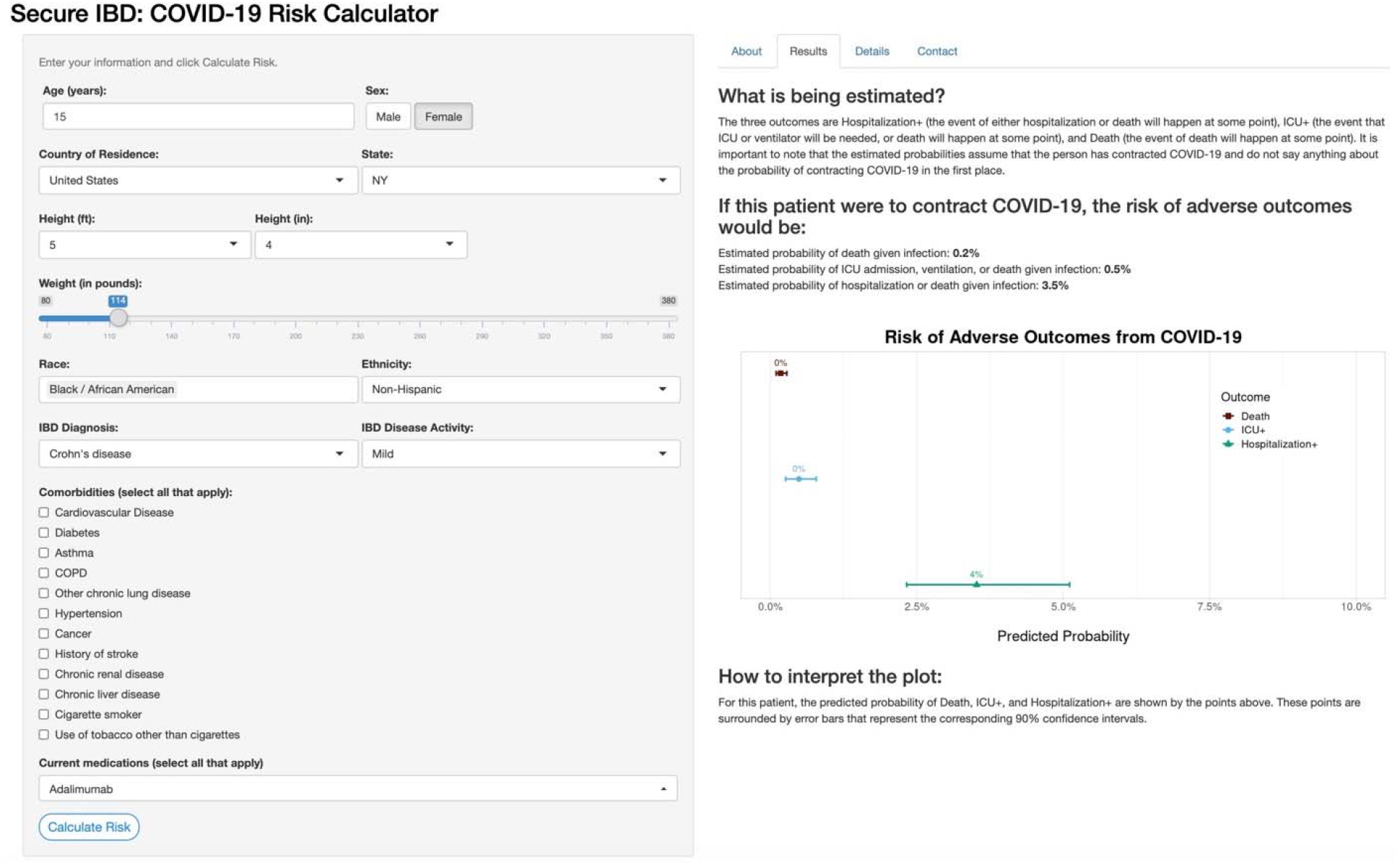
An example patient with below-average predicted risk. Young age, gender, and a lack of comorbidities contribute to a lower-than-average risk of adverse COVID-19 outcomes.

**Figure 3B.**
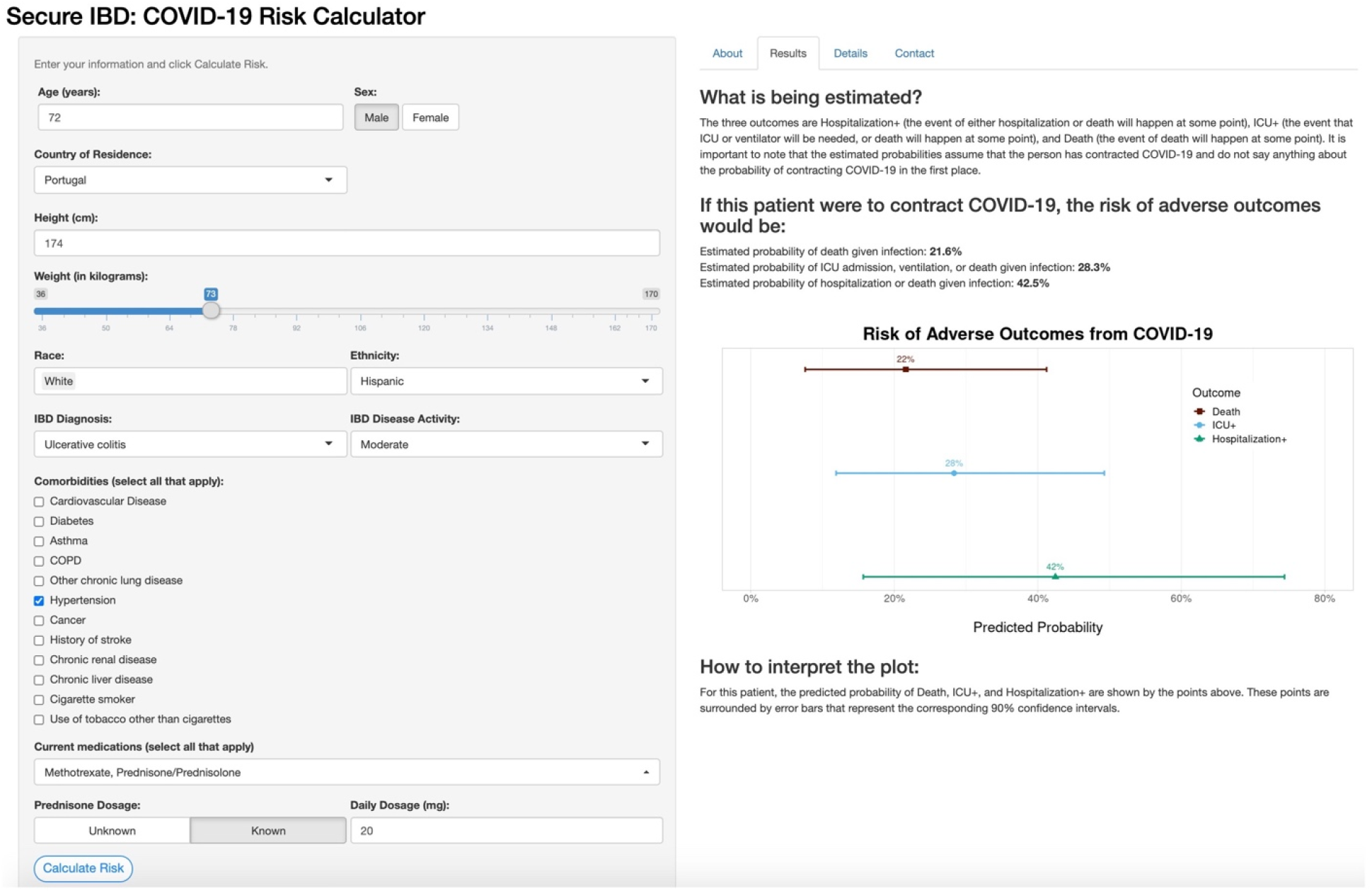
An example patient with above-average predicted risk. Older age, prednisone dosage, and hypertension were the major contributors to increased risk, with ethnicity having a small positive association with increased risk. Figure 3: Online Risk Prediction Tool. The data entry and results for two example patients are depicted.

## Discussion

We developed and validated risk prediction models for hospitalization, intensive care stay, and death resulting from COVID-19 in patients with IBD using data from 2,709 cases from 59 countries reported through an international voluntary registry. ^13^. The models demonstrated excellent discrimination, with a test set AUC (95% CI) of 0.79 (0.75, 0.83) for Hospitalization+, 0.88 (0.82, 0.95) for ICU+, and 0.94 (0.89, 0.99) for Death. We made a free online risk calculator utilizing these models (https://covidibd.org/covid-19-risk-calculator/) for healthcare providers to facilitate discussion of risks due to COVID-19 with their IBD patients^1^. The interactive application could provide a reliable basis for distinguishing between high- and low-risk patients to aid in personalizing clinical guidance on decisions about precautions, returning to normal activities, and vaccination. Other COVID-19-related risk prediction tools have focused on predicting hospital course based on clinical data captured at the time of admission^9^ and predicting mortality among US patients aged 65 years and older^8^. Our risk tool is unique in at least three ways. First, we focus on a specialized population of patients with IBD, a chronic, immune-mediated condition frequently treated with immune suppressive medications and often affected by other comorbidities. Second, our model focuses on predictors that are known before a patient were to contract COVID-19 and thus can be used to inform lifestyle or treatment decisions to prevent infection or downstream complications. Finally, we examine a broader range of outcomes than tools focused solely on mortality. Our work can serve as a model for other disease areas, and our code is publicly available and could be adapted for similar online risk tools in other settings or populations.

Strong associations with worse adverse COVID-19 outcomes were oral corticosteroids, older age, comorbidities, gender, and white physician-reported race (for Hospitalization+). Caution must be used when interpreting penalized regression results because the coefficients are biased, but the results for oral corticosteroids were particularly dramatic. Compared to not taking an oral corticosteroid, taking a daily dose equivalent of 40mg of prednisone was associated with a ten times greater adjusted odds of death. Biologic therapies were associated with a lower risk of adverse COVID-19 outcomes, with small differences between the subcategories of biologic therapies. Compared to not taking a biologic therapy, TNF inhibitors were associated with an adjusted odds ratio of 0.62 for death. In contrast to earlier studies using this database^14,32^, we did not find a consistent association between 5-aminosalicylates and a higher risk of adverse outcomes; depending on the imputation and bootstrap replication, the sign would often change from positive to negative.

The worldwide collaboration that enabled this study and the detailed clinical data reported by physicians or trained medical staff is an important strength of this study. The machine learning approach allowed us to consider a wide variety of potential associations with adverse outcomes, and we examined multiple adverse COVID-19-related adverse outcomes enabling preliminary comparisons between risks. Certain comorbidities, including COPD, CVD, and cancer, were not as strongly associated with Hospitalization+ as they were for death. In contrast, severe IBD disease activity was an important predictor of Hospitalization+ but was not consistently associated with a higher risk of ICU+ or death.

### Limitations

The data for this study comes from a voluntary registry, and the sampling process is unknown. Reported cases may underrepresent both low-risk asymptomatic cases and severely ill patients who may be hospitalized at an outside hospital or die without their healthcare provider’s knowledge. Our results are associational, not causal — when using the online risk calculator, healthcare providers should not use it to answer “what-if” questions (e.g., how an individual’s risk would change if they altered the medications they were taking) which are inherently causal questions^33^. While the registry has a wealth of clinical data, it does not collect granular data on many social determinants of health. Additionally, insurance status is not collected, which, for patients in the US, likely factors into the decision making of whether to visit a hospital. Lastly, we cannot compare the risk of adverse COVID-19 outcomes in IBD patients to that in the general population.

## Conclusions

This prognostic model can effectively predict which IBD patients may be at higher risk for COVID-19-related morbidity. The free and publicly available (https://covidibd.org/covid-19-risk-calculator/) risk calculator should facilitate patient-provider discussions regarding the individualized risk of COVID-19 based on patient and treatment-related factors. As COVID-19 cases continue to rise in the US and the rest of the world, this tool will be important in assisting physicians in identifying high-risk patients with downstream clinical implications. This tool can inform public health efforts to promote rational vaccine allocation and could help providers target their outreach to higher-risk patients. We believe this approach can also serve as a model for risk stratification in other chronic diseases.

## Supporting information

TRIPOD Checklist

## Data Availability

We are committed to sharing our data with the international research community. Data requests are reviewed by our SECURE-IBD team including the International Advisory Committee to ensure data will be used in a scientifically and ethically sound way. The data request form and additional information can be found online at https://covidibd.org/sharing-secure-ibd-data/

## Acknowledgments

This work was funded by the Helmsley Charitable Trust (2003-04445), National Center for Advancing Translational Sciences (UL1TR002489), a T32DK007634 (EJB), and a K23KD111995-01A1 (RCU). Additional funding was provided by Pfizer, Takeda, Janssen, Abbvie, Eli Lilly, Genentech, Boehringer Ingelheim, Bristol Myers Squibb, Celltrion, and Arenapharm.

## Conflicts of Interest

John Sperger, Kushal S. Shah, Minxin Lu, Xian Zhang, Erica J. Brenner, Manasi Agrawal, and Michael R. Kosorok report no conflicts of interest.

RCU has served as a consultant and/or advisory board member for Bristol Myers Squibb, Eli Lilly, Janssen, Pfizer, and Takeda. He has received research support from AbbVie, Boehringer Ingelheim, and Pfizer. He is supported by a Career Development Award from the National Institutes of Health (K23KD111995-01A1).

JFC reports receiving research grants from AbbVie, Janssen Pharmaceuticals and Takeda; receiving payment for lectures from AbbVie, Amgen, Allergan, Inc. Ferring Pharmaceuticals, Shire, and Takeda; receiving consulting fees from AbbVie, Amgen, Arena Pharmaceuticals, Boehringer Ingelheim, Celgene Corporation, Celltrion, Eli Lilly, Enterome, Ferring Pharmaceuticals, Genentech, Janssen Pharmaceuticals, Landos, Ipsen, Medimmune, Merck, Novartis, Pfizer, Shire, Takeda, Tigenix, Viela bio; and hold stock options in Intestinal Biotech Development and Genfit.

MDK has consulted for Abbvie, Janssen, Pfizer, and Takeda, is a shareholder in Johnson & Johnson, and has received research support from Pfizer, Takeda, Janssen, Abbvie, Lilly, Genentech, Boehringer Ingelheim, Bristol Myers Squibb, Celtrion, and Arenapharm.

## Notes

### Author Declarations

The UNC-Chapel Hill Office for Ethics has determined that the storage and analysis of de-identified data do not constitute human subject research (SOP 4201 https://unc.policystat.com/policy/5160241/latest/) as defined under [45 CFR 46.102 and 21 CFR 56.102] and do not require IRB approval.

