## Supplemental tables and figures for "Development and validation of multivariable prediction models for adverse COVID-19 outcomes in IBD patients"

eTable 1. Variables included in the model

eTable 2. Characteristics of COVID-19 IBD Patients in the Study

eFigure 1: Estimated Contrast Sign Distribution

eFigure 2: Estimated Contrast Effect Size Distribution

eFigure 3: Estimated Country Sign Distribution

eFigure 4: Estimated Country Effect Size Distribution

eFigure 5: Estimated U.S. Region Sign Distribution

eFigure 6: Estimated U.S. Region Effect Size Distribution

References

**eTable 1.** **Variables included in the model**

| **Domain** | **Name of Variable** | **Scale** | **Description of the variable** |
| --- | --- | --- | --- |
| Outcome variables | Hospitalization+ | B | A composite outcome of hospitalization, ICU admission, mechanical ventilation, or death |
|  | ICU+ | B | A composite outcome of ICU admission, mechanical ventilation, or death |
|  | Death | B | Indicator for outcome of death |
| Predictors | Age | I | Age in years. Age is capped at 90 |
|  | Date | I | Date of report submission. |
|  | Gender | C | Male or female gender |
|  | Race | M | Physician-reported race. Three categories were included, Asian, Black, and White. were included as binary variables. Other race categories have prevalence lower than 30 reports. One individual can be assigned to more than one race category. |
|  | Ethnicity | B | Indicator of whether the person is Hispanic or Latino |
|  | Height | X | Height in cm |
|  | BMI | X | Body Mass Index = weight (kg) / (height (m))^2^  Treated as missing if BMIs are above 60 |
|  | Weight | X | Weight in kg. Treated as missing if BMI was above 60 or |
|  | Country | C | Countries with less than 30 entries in our dataset are grouped to level “others” |
|  | Country income level | C | World bank income classification^1^ . Low and lower-middle income countries were collapsed together due to the small number of reports from low-income countries. |
|  | US Census Division | C | Place of residence in the US according to US census division, except for New York which gets its own indicator. Individuals who reside in other countries are assigned to the category “Other county” which is excluded from the model because it is redundant with the Country variable. |
|  | Disease type | C | Crohn's disease or Ulcerative colitis |
|  | Disease activity | C | Includes: remission, mild, moderate, severe. Each of the activity category is recorded as a binary variable. |
|  | Comorbidities | M | Includes: asthma, cancer, cardiovascular disease, chronic liver disease, chronic renal disease, COPD, other chronic lung disease (not Asthma/COPD), diabetes, hypertension, history of stroke and current smoker. Each of the comorbidity is recorded as a binary variable. |
|  | Count of comorbidities | N | Number of comorbidities entered |
|  | Medication category | M | IBD medications at time of COVID diagnosis, which Includes: biologic therapy, 5-aminosalicylates, immunomodulators, corticosteroids, Janus kinase inhibitors (tofacitinib), other IBD medication, and no IBD medication. Each of the category is recorded as a binary variable. One participant can take multiple medications. |
|  | Sulfasalazine | B | Indicator for the use of Sulfasalazine |
|  | Mesalamine | B | Indicator for the use of Mesalamine |

**eTable 1. Variables included in the model (continued)**

| **Domain** | **Name of Variable** | **Scale** | **Description of the variable** |
| --- | --- | --- | --- |
| Predictors | Azathioprine daily dose | X | Total azathioprine dosage in mg per day |
|  | 6-Mercaptopurine daily dose | X | Total 6-mercaptopurine dosage in mg per day |
|  | Prednisone daily dose | X | Effective daily prednisone dosage in mg. Different medication frequencies were translated into a daily equivalent. |
|  | Tumor Necrosis Factor inhibitor | B | Indicator for the use of Tumor Necrosis Factor inhibitor |
|  | Budesonide | B | Indicator for the use of Budesonide |
|  | Oral or parenteral steroids | B | Indicator for the use of Oral or parenteral steroids |
|  | Anti-integrin | B | Indicator for the use of Anti-integrin |
|  | IL 12/23 inhibitor | B | Indicator for the use of IL 12/23 inhibitor |
|  | Immunomodulator: Methotrexate | B | Indicator for the use of Methotrexate as an immunomodulator |
|  | Immunomodulator: Azathioprine or 6-Mercaptopurine | B | Indicator for the use of Azathioprine or 6-Mercaptopurine as an immunomodulator |
| Transformations | Azathioprine daily dose by weight | X | Azathioprine daily dose divided by weight, in the unit of mg per day per kg |
|  | 6-Mercaptopurine daily dose by weight | X | 6-Mercaptopurine daily dose divided by weight, in the unit of mg per day per kg |
|  | Prednisone daily dose by weight | X | Prednisone daily dose divided by weight, in the unit of mg per day per kg |
|  | Age^2 | X | Quadratic term of age |
|  | Count of comorbidities ^2 | I | Quadratic term of the count of comorbidities |
|  | BMI^2 | X | Quadratic term of BMI |
|  | Prednisone daily dose ^2 | X | Quadratic term of prednisone daily dose |
|  | Azathioprine daily dose ^2 | X | Quadratic term of azathioprine daily dose |
|  | 6-Mercaptopurine daily dose ^2 | X | Quadratic term of 6-mercaptopurine daily dose |
| Interaction terms | Age: Gender | X | Interaction term between age and gender |
|  | Age: Count of comorbidities | X | Interaction term between age and Count of comorbidities |
|  | Age: BMI | X | Interaction term between age and BMI |
|  | Date:Country | X | Interaction term between Date Interval and country |
|  | Cardiovascular disease: Diabetes | B | Interaction term between Cardiovascular disease and Diabetes |
|  | Cardiovascular disease: Hypertension | B | Interaction term between Cardiovascular disease and Hypertension |
|  | Diabetes: Hypertension | B | Interaction term between Diabetes and Hypertension |
|  | Hypertension: Chronic renal disease | B | Interaction term between Hypertension and Chronic renal disease |
|  | Current smoker: Count of comorbidities | N | Interaction term between Current smoker and Count of comorbidities |

**eTable 1. Variables included in the model (continued)**

| **Domain** | **Name of Variable** | **Scale** | **Description of the variable** |
| --- | --- | --- | --- |
| Interaction terms | Biologic therapy: 5-aminosalicylates | B | Interaction term between Biologic therapy and 5-aminosalicylates |
|  | Biologic therapy: Immunomodulators | B | Interaction term between Biologic therapy and Immunomodulators |
|  | Biologic therapy: Corticosteroids | B | Interaction term between Biologic therapy and Corticosteroids |
|  | 5-aminosalicylates: Immunomodulators | B | Interaction term between 5-aminosalicylates and Immunomodulators |
|  | 5-aminosalicylates: Corticosteroids | B | Interaction term between 5-aminosalicylates and Corticosteroids |
|  | Immunomodulators: Corticosteroids | B | Interaction term between Immunomodulators and Corticosteroids |
|  | Oral/parenteral steroids: Age | X | Interaction term between Oral/parenteral steroids and age |
|  | Oral/parenteral steroids: Gender | B | Interaction term between Oral/parenteral steroids and gender |
|  | Oral/parenteral steroids: BMI | X | Interaction term between Oral/parenteral steroids and BMI |

Scales: B = binary, C = categorical, I = integers, M = multidimensional, N = count, X = continuous.
 ^1^world bank classification of country by income

**eTable 2. Characteristics of COVID-19 IBD Patients in the Study**

| **Characteristics** | **Training Data (n=2009)** | **Test Data (n=700)** | **Overall (n=2709)** |
| --- | --- | --- | --- |
| Age, mean (SD), years | 42.2 (18.2) | 38.7 (17.4) | 41.2 (18.0) |
| Missing | 9 (0.4%) | 4 (0.6%) | 13 (0.5%) |
| Gender, No. (%) |  |  |  |
| Female | 982 (48.9%) | 344 (49.1%) | 1326 (48.9%) |
| Male | 998 (49.7%) | 341 (48.7%) | 1339 (49.4%) |
| Other | 1 (0.0%) | 0 (0.0%) | 1 (0.0%) |
| Missing | 28 (1.4%) | 15 (2.1%) | 43 (1.6%) |
| Asian*, No. (%) | 112 (5.6%) | 38 (5.4%) | 150 (5.5%) |
| Black*, No. (%) | 138 (6.9%) | 39 (5.6%) | 177 (6.5%) |
| White*, No. (%) | 1603 (79.8%) | 547 (78.1%) | 2150 (79.4%) |
| Hispanic/Latino, No. (%) | 350 (17.4%) | 115 (16.4%) | 465 (17.2%) |
| Missing | 375 (18.7%) | 120 (17.1%) | 495 (18.3%) |
| Month of entry, No. (%) |  |  |  |
| March | 195 (9.7%) | 32 (4.6%) | 227 (8.4%) |
| April | 567 (28.2%) | 94 (13.4%) | 661 (24.4%) |
| May | 391 (19.5%) | 78 (11.1%) | 469 (17.3%) |
| June | 232 (11.5%) | 47 (6.7%) | 279 (10.3%) |
| July | 258 (12.8%) | 49 (7.0%) | 307 (11.3%) |
| August | 238 (11.8%) | 35 (5.0%) | 273 (10.1%) |
| September | 128 (6.4%) | 180 (25.7%) | 308 (11.4%) |
| October | 0 (0%) | 185 (26.4%) | 185 (6.8%) |
| Height, mean (SD), cm | 169 (10.7) | 169 (12.0) | 169 (11.1) |
| Missing | 375 (18.7%) | 102 (14.6%) | 477 (17.6%) |
| BMI, mean (SD) | 26.0 (6.50) | 25.2 (6.26) | 25.8 (6.44) |
| Missing | 398 (19.8%) | 106 (15.1%) | 504 (18.6%) |
| Weight, mean (SD), kg | 74.7 (20.9) | 72.4 (20.8) | 74.1 (20.9) |
| Missing | 398 (19.8%) | 106 (15.1%) | 504 (18.6%) |
| Country, No. (%) |  |  |  |
| Argentina | 38 (1.9%) | 23 (3.3%) | 61 (2.3%) |
| Belgium | 43 (2.1%) | 16 (2.3%) | 59 (2.2%) |
| Brazil | 65 (3.2%) | 24 (3.4%) | 89 (3.3%) |
| Canada | 27 (1.3%) | 11 (1.6%) | 38 (1.4%) |
| France | 95 (4.7%) | 20 (2.9%) | 115 (4.2%) |
| Germany | 38 (1.9%) | 12 (1.7%) | 50 (1.8%) |
| India | 23 (1.1%) | 10 (1.4%) | 33 (1.2%) |
| Iran, Islamic Republic of | 55 (2.7%) | 5 (0.7%) | 60 (2.2%) |
| Israel | 25 (1.2%) | 12 (1.7%) | 37 (1.4%) |
| Italy | 74 (3.7%) | 18 (2.6%) | 92 (3.4%) |
| Netherlands | 27 (1.3%) | 21 (3.0%) | 48 (1.8%) |
| Portugal | 22 (1.1%) | 10 (1.4%) | 32 (1.2%) |
| Russian Federation | 119 (5.9%) | 46 (6.6%) | 165 (6.1%) |
| Spain | 235 (11.7%) | 53 (7.6%) | 288 (10.6%) |
| Turkey | 30 (1.5%) | 20 (2.9%) | 50 (1.8%) |
| United Kingdom | 94 (4.7%) | 29 (4.1%) | 123 (4.5%) |
| United States | 800 (39.8%) | 276 (39.4%) | 1076 (39.7%) |
| Others^†^ | 199 (9.9%) | 94 (13.4%) | 293 (10.8%) |

**eTable 2. Characteristics of COVID-19 IBD Patients in the Study (continued)**

| **Characteristics** | **Training Data (n=2009)** | **Test Data (n=700)** | **Overall (n=2709)** |
| --- | --- | --- | --- |
| Country income level, No. (%) |  |  |  |
| Low or lower-middle | 26 (1.3%) | 14 (2.0%) | 40 (1.5%) |
| Upper-middle | 360 (17.9%) | 134 (19.1%) | 494 (18.2%) |
| High | 1623 (80.8%) | 545 (77.9%) | 2168 (80.0%) |
| Missing | 0 (0%) | 7 (1.0%) | 7 (0.3%) |
| US Census Division*, No. (%) |  |  |  |
| East North Central | 119 (5.9%) | 38 (5.4%) | 157 (5.8%) |
| East South Central | 60 (3.0%) | 27 (3.9%) | 87 (3.2%) |
| West | 70 (3.5%) | 28 (4.0%) | 98 (3.6%) |
| West North Central | 50 (2.5%) | 28 (4.0%) | 78 (2.9%) |
| West South Central | 77 (3.8%) | 21 (3.0%) | 98 (3.6%) |
| New England | 61 (3.0%) | 11 (1.6%) | 72 (2.7%) |
| Mid-Atlantic | 64 (3.2%) | 22 (3.1%) | 86 (3.2%) |
| South Atlantic | 117 (5.8%) | 50 (7.1%) | 167 (6.2%) |
| New York | 181 (9.0%) | 47 (6.7%) | 228 (8.4%) |
| Other country | 1209 (60.2%) | 421 (60.1%) | 1630 (60.2%) |
| Missing | 1 (0.0%) | 7 (1.0%) | 8 (0.3%) |
| Current smoker, No. (%) | 61 (3.0%) | 25 (3.6%) | 86 (3.2%) |
| Disease type, No. (%) |  |  |  |
| Crohn's disease | 1115 (55.5%) | 401 (57.3%) | 1516 (56.0%) |
| Ulcerative colitis | 854 (42.5%) | 278 (39.7%) | 1132 (41.8%) |
| Missing | 40 (2.0%) | 21 (3.0%) | 61 (2.3%) |
| Disease activity |  |  |  |
| Remission, No. (%) | 1099 (54.7%) | 411 (58.7%) | 1510 (55.7%) |
| Mild, No. (%) | 394 (19.6%) | 137 (19.6%) | 531 (19.6%) |
| Moderate, No. (%) | 316 (15.7%) | 101 (14.4%) | 417 (15.4%) |
| Severe, No. (%) | 118 (5.9%) | 26 (3.7%) | 144 (5.3%) |
| Missing, No. (%) | 82 (4.1%) | 25 (3.6%) | 107 (3.9%) |
| Asthma, No. (%) | 111 (5.5%) | 21 (3.0%) | 132 (4.9%) |
| Cancer, No. (%) | 41 (2.0%) | 9 (1.3%) | 50 (1.8%) |
| Cardiovascular disease, No. (%) | 133 (6.6%) | 43 (6.1%) | 176 (6.5%) |
| Chronic liver disease, No. (%) | 68 (3.4%) | 27 (3.9%) | 95 (3.5%) |
| Chronic renal disease, No. (%) | 44 (2.2%) | 14 (2.0%) | 58 (2.1%) |
| COPD, No. (%) | 33 (1.6%) | 16 (2.3%) | 49 (1.8%) |
| Other chronic lung disease (not Asthma/COPD), No. (%) | 31 (1.5%) | 7 (1.0%) | 38 (1.4%) |
| Diabetes, No. (%) | 117 (5.8%) | 30 (4.3%) | 147 (5.4%) |
| Hypertension, No. (%) | 243 (12.1%) | 67 (9.6%) | 310 (11.4%) |
| History of stroke, No. (%) | 26 (1.3%) | 8 (1.1%) | 34 (1.3%) |
| Count of comorbidities, mean (SD) | 0.544 (0.938) | 0.479 (0.879) | 0.527 (0.923) |
| Biologic therapy, No. (%) | 1203 (59.9%) | 437 (62.4%) | 1640 (60.5%) |
| Tumor Necrosis Factor inhibitor, No. (%) | 796 (39.6%) | 313 (44.7%) | 1109 (40.9%) |
| Anti-integrin, No. (%) | 213 (10.6%) | 72 (10.3%) | 285 (10.5%) |
| IL 12/23 inhibitor, No. (%) | 187 (9.3%) | 47 (6.7%) | 234 (8.6%) |
| 5-Aminosalicylates, No. (%) | 636 (31.7%) | 200 (28.6%) | 836 (30.9%) |
| Sulfasalazine, No. (%) | 64 (3.2%) | 24 (3.4%) | 88 (3.2%) |
| Mesalamine, No. (%) | 561 (27.9%) | 175 (25.0%) | 736 (27.2%) |
| Immunomodulators, No. (%) | 459 (22.8%) | 140 (20.0%) | 599 (22.1%) |

**eTable 2. Characteristics of COVID-19 IBD Patients in the Study (continued)**

| **Characteristics** | **Training Data (n=2009)** | **Test Data (n=700)** | **Overall (n=2709)** |
| --- | --- | --- | --- |
| Methotrexate, No. (%) | 81 (4.0%) | 24 (3.4%) | 105 (3.9%) |
| Azathioprine or 6-Mercaptopurine, No. (%) | 367 (18.3%) | 110 (15.7%) | 477 (17.6%) |
| Azathioprine dose among takers, mean (SD), mg | 116.8 (49.2) | 115.9 (51.6) | 116.6 (49.6) |
| Missing | 16 (0.8%) | 5 (0.7%) | 21 (0.8%) |
| 6-Mercaptopurine dose among takers, mean (SD), mg | 61.5 (24.8) | 60.0 (24.9) | 61.0 (24.7) |
| Missing | 1 (0.0%) | 3 (0.4%) | 4 (0.1%) |
| Corticosteroids, No. (%) | 209 (10.4%) | 65 (9.3%) | 274 (10.1%) |
| Budesonide, No. (%) | 59 (2.9%) | 17 (2.4%) | 76 (2.8%) |
| Prednisone daily dose among takers, mean (SD), mg/day | 26.8 (18.3) | 22.7 (15.2) | 25.8 (17.6) |
| Missing | 76 (3.8%) | 20 (2.9%) | 96 (3.5%) |
| Oral or parenteral steroids, No. (%) | 154 (7.7%) | 49 (7.0%) | 203 (7.5%) |
| Janus kinase inhibitors (tofacitinib), No. (%) | 30 (1.5%) | 9 (1.3%) | 39 (1.4%) |
| Other IBD medication, No. (%) | 63 (3.1%) | 23 (3.3%) | 86 (3.2%) |
| No IBD medication, No. (%) | 70 (3.5%) | 25 (3.6%) | 95 (3.5%) |
| Hospitalized, No. (%) | 497 (24.7%) | 136 (19.4%) | 633 (23.4%) |
| Missing | 27 (1.3%) | 11 (1.6%) | 38 (1.4%) |
| Ventilator, No. (%) | 74 (3.7%) | 17 (2.4%) | 91 (3.4%) |
| Missing | 34 (1.7%) | 14 (2.0%) | 48 (1.8%) |
| ICU, No. (%) | 95 (4.7%) | 28 (4.0%) | 123 (4.5%) |
| Missing | 38 (1.9%) | 13 (1.9%) | 51 (1.9%) |
| Hospitalization+, No. (%) | 499 (24.8%) | 138 (19.7%) | 637 (23.5%) |
| Missing | 37 (1.8%) | 17 (2.4%) | 54 (2.0%) |
| ICU+, No. (%) | 133 (6.6%) | 36 (5.1%) | 169 (6.2%) |
| Missing | 49 (2.4%) | 22 (3.1%) | 71 (2.6%) |
| Death, No. (%) | 57 (2.8%) | 12 (1.7%) | 69 (2.5%) |
| Missing | 33 (1.6%) | 14 (2.0%) | 47 (1.7%) |

* Individuals can be assigned to more than one physician-reported racial group (White, Black, Asian).
^†^ Countries with fewer than 30 submissions were grouped together into the Other countries category

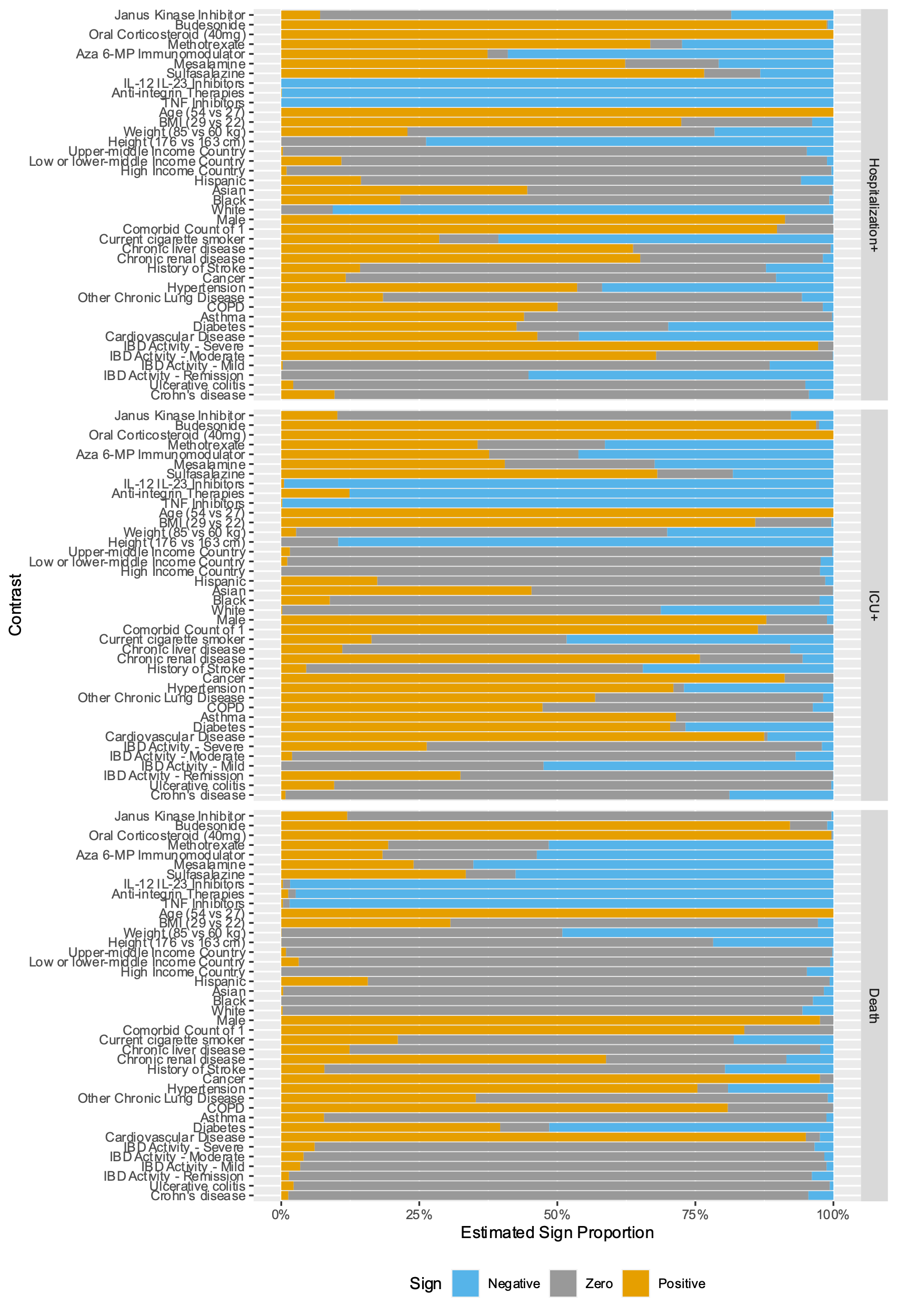

**eFigure 1: Estimated Contrast Sign Distribution**

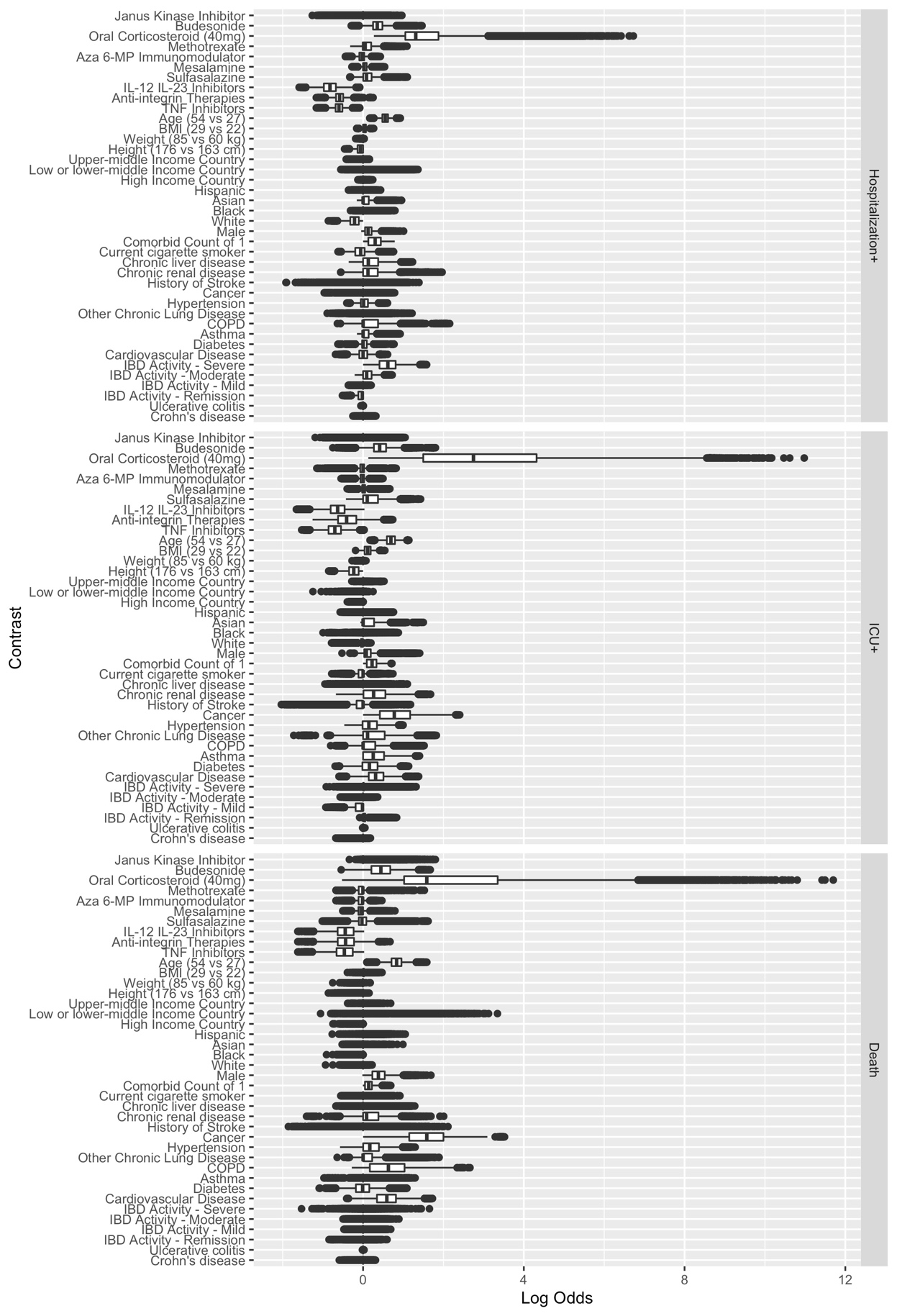

**eFigure 2: Estimated Contrast Effect Size Distribution**

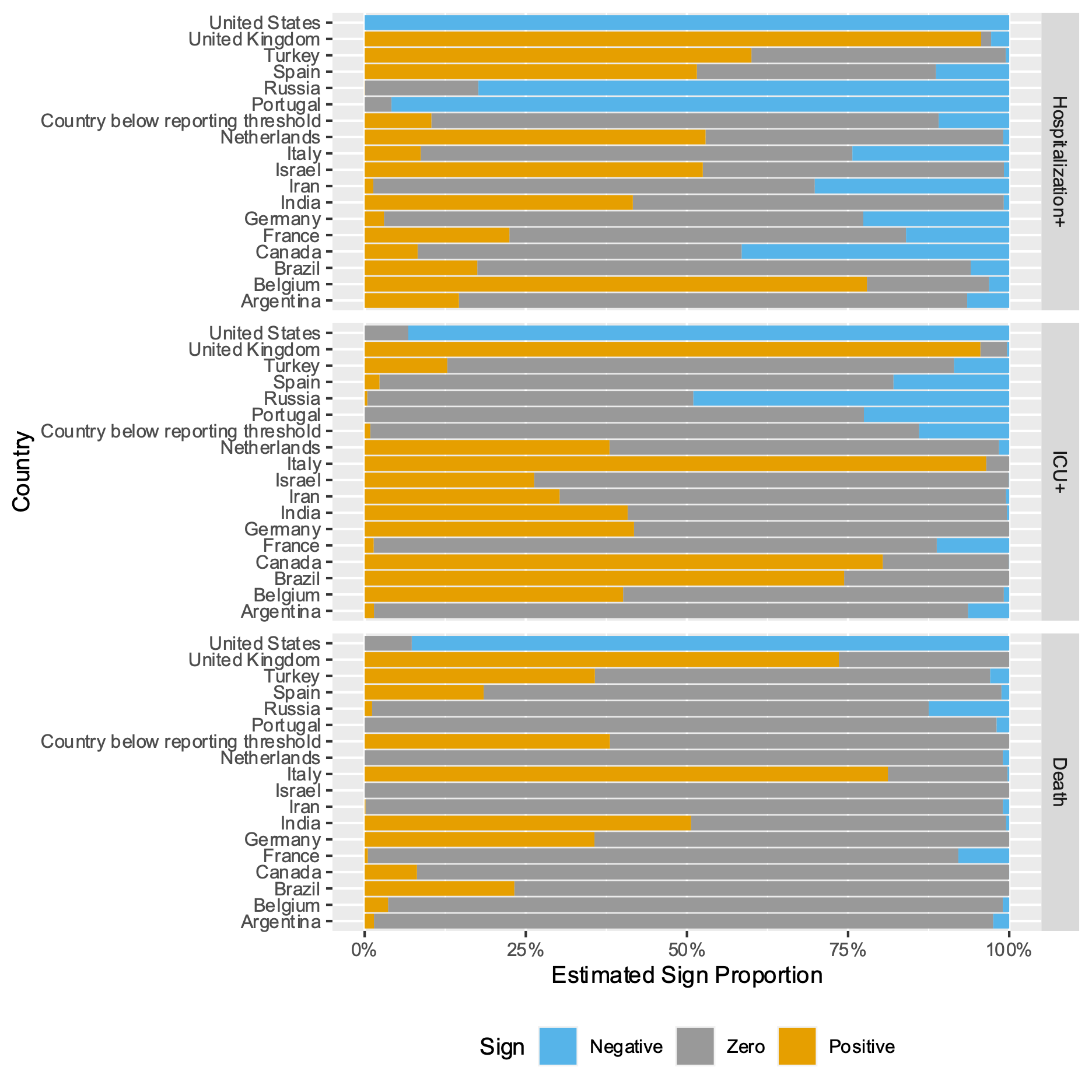

**eFigure 3: Estimated Country Sign Distribution**

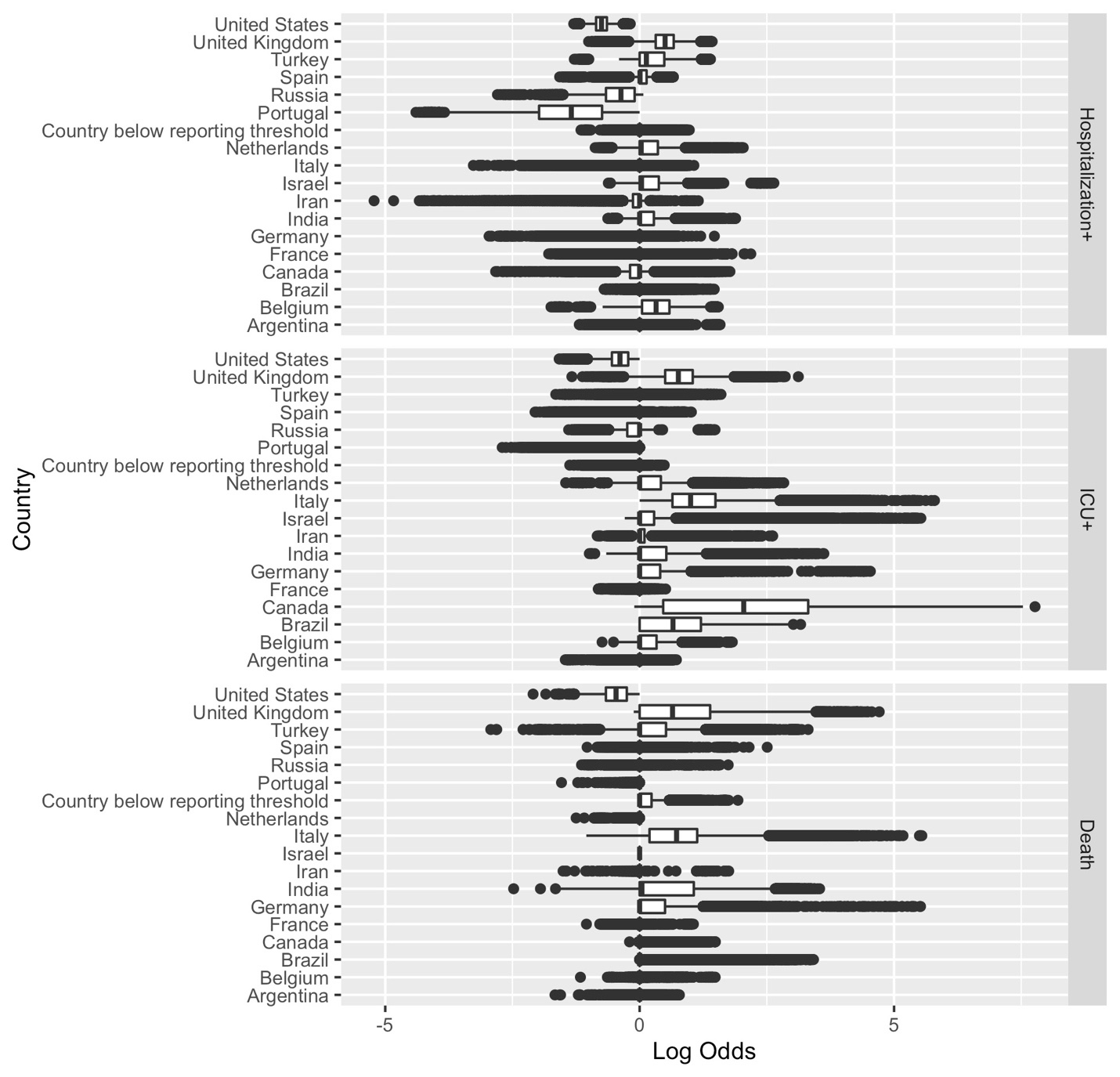

**eFigure 4: Estimated Country Effect Size Distribution**

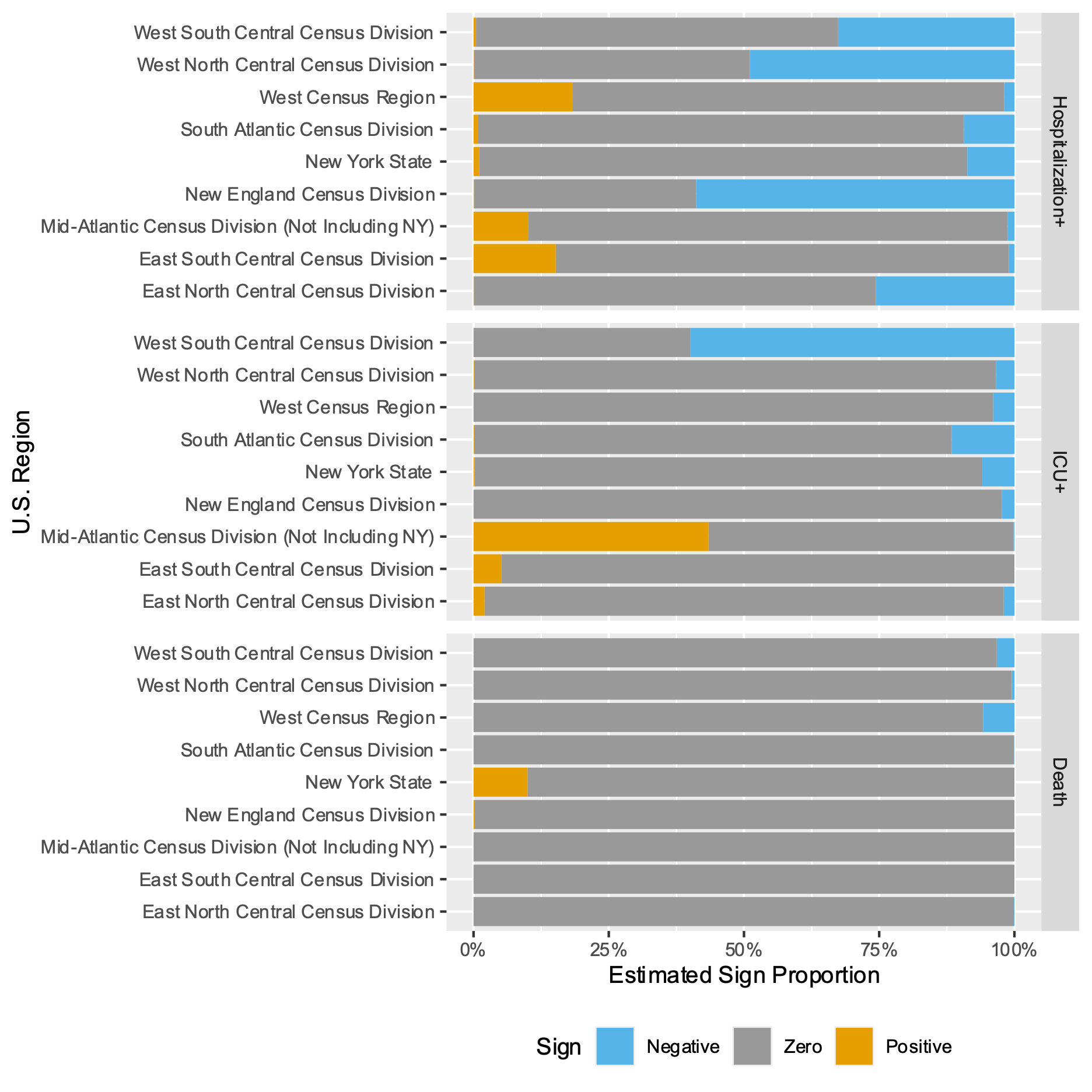

**eFigure 5: Estimated U.S. Region Sign Distribution**

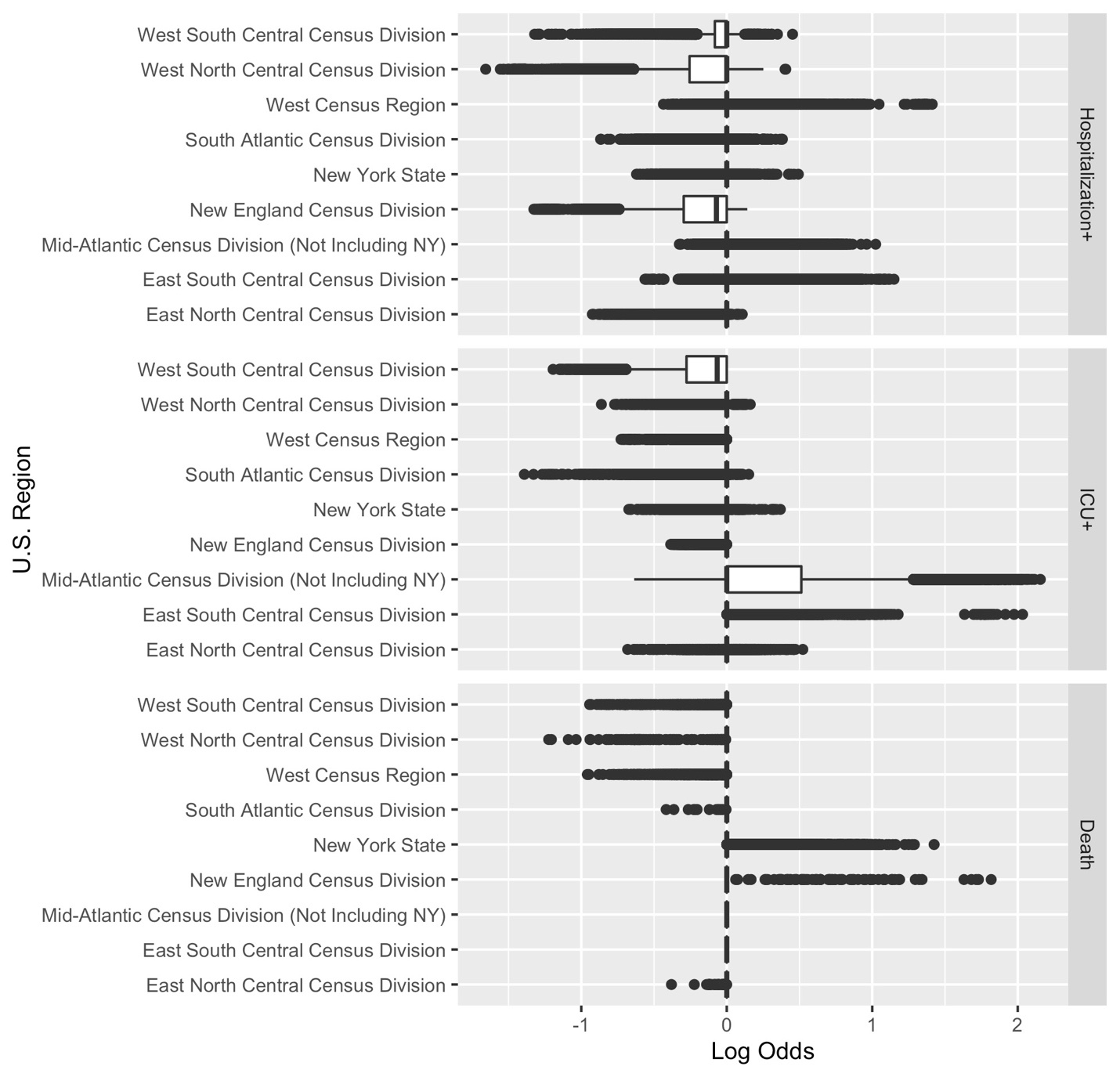

**eFigure 6: Estimated U.S. Region Effect Size Distribution**
